# Surveying Tenants of Permanent Supportive Housing in Skid Row about COVID-19

**DOI:** 10.1101/2020.04.17.20070052

**Authors:** Benjamin F. Henwood, Brian Redline, Jack Lahey

## Abstract

Permanent supportive housing (PSH) enrolls highly vulnerable homeless adults who experience early onset of geriatric conditions and require in-home support. Thus, there is potentially a high risk for COVID-19 within PSH, which may necessitate tenants to take protective measures. This study reports on survey results collected from 532 PSH tenants in Los Angeles, California during the 4^th^ week of March in 2020. Results show that nearly all tenants were aware of COVID-19, and 65% considered it to be a very serious health threat. The latter characteristic was a strong predictor of taking protective measures (i.e., handwashing and social distancing). Tenants in units with shared bathroom facilities had lower odds of social distancing than those in studio apartments. Tenants with mental health diagnoses had lower odds of consistent handwashing. Lack of access to food, hygiene items, and medication delivery were commonly reported barriers to sheltering in place.

---

More than half a million adults are homeless in the United States,^1^ and these individuals constitute a high-risk group for the novel coronavirus disease 2019 (COVID-19). Living in shelters or on the streets makes protective measures including social distancing and handwashing difficult. In addition, high rates of underlying health conditions in the homeless population,^2^ including obstructive lung disease,^3^ increase vulnerability.^4^ An estimated 21,000 people experiencing homelessness in the United States could require hospitalization at a peak infection rate of 40% among this population.^5^ As a matter of policy, the most vulnerable homeless adults are prioritized for Housing First programs,^6^ which means they receive immediate access to independent living situations coupled with support services—also known as permanent supportive housing (PSH).^7^ In 2019, the United States had more than 369,000 PSH units^1^ capable of providing opportunities for social distancing. Older units such as single-room occupancy (SRO) facilities with shared bathroom facilities, however, may make social distancing more challenging.

Whether PSH tenants are aware of COVID-19 and are taking protective measures is largely unknown. Among the general population, engagement in social distancing and handwashing was most strongly predicted by the perceived likelihood of being infected.^8^ Based on surveys conducted during the first official week of the pandemic (March 11-16, 2020) among the general public regarding perceptions and responses to the novel coronavirus epidemic, most of the general population considers the current outbreak to be serious and is taking protective actions including increased handwashing and social distancing. However, only about half of respondents who experience symptoms of the virus (e.g. fever or chills and shortness of breath) take steps to stay away from other people.^9^ Also concerning, the surveys demonstrated that there are subgroups that are largely disengaged, unaware, and not practicing protective behaviors.^8^

This study reports on survey results collected from PSH tenants in the Skid Row and downtown areas of Los Angeles during the week (March 20–27, 2020) after California’s governor issued a statewide shelter-in-place order.^10^ This area represents a concentrated area of homelessness where the risk of exposure and spread is high due to close contact of people living on the streets and lack of sanitation. In this study we examine tenants’ knowledge of COVID-19, perceived risk, pre-existing condition risk factors, consistency of handwashing and social distancing since the outbreak began, recent experiences of flu-like symptoms, and tenants’ ability to shelter in place. A better understanding of these COVID-19-related factors among PSH tenants will serve to inform policies and interventions to be implemented by service providers in order to protect this vulnerable population.

## Methods

Staff members from one of the largest providers of PSH in Los Angeles conducted phone surveys of tenants who lived in either a studio apartment or an SRO with shared bathroom facilities. Tenant responses were entered into a survey tool and analyzed to guide programmatic response to tenant needs. For this report, tenant responses were merged with demographic characteristics from existing administration records, deidentified, and analyzed for research purposes. Approximately half of all tenants were surveyed (n=766), with PSH staff reporting no refusals. Of these, 532 were successfully matched to complete demographic data and included for analysis; respondents in our analytic sample were on average 1.5 years older than tenants with demographic data who did not respond (*p* =.02). The study was approved as exempt by the first author’s University Institutional Review Board.

### Measures

Surveys began by asking residents if they had “heard of the coronavirus/COVID-19 outbreak (yes; no),” and they were provided information about it if not. The following questions concerned how they viewed the health risk posed by COVID-19 (“Do you view this outbreak as a serious risk to your health? [not at all; not very seriously; somewhat seriously; very seriously]”); whether they had a pre-existing condition that puts them in a high risk group for COVID-19 (“Are you in a category considered to be at serious risk for coronavirus/COVID-19? [I have a preexisting condition; I am over 60 years old; I’m not sure; no]”); whether they had consistently been engaging in preventive handwashing (“have you been washing your hands more since the outbreak began? [no; yes, once in while; yes, sometimes; yes, all the time]”) and social distancing (“have you been social distancing, or staying away from others and limiting social gatherings, to avoid spreading the virus? [no; yes, once in while; yes, sometimes; yes, all the time]”); and whether they had experienced flu-like symptoms in the prior 30 days (“Have you been sick in the past 30 days with cold or flu-like symptoms? [no; yes – cold only; yes – flu only; yes – both]”). Tenants were also asked whether they could shelter in place for 14 days if necessary (“Are you able to shelter in place (i.e. not leave your home and practice social distancing at all times) for 14 days? [no; maybe; yes]”), and those who answered “no” or “maybe” were asked, “If not, what would you need so that you can shelter in place that you are not able to secure for yourself? [select all that apply: delivery of food; hygiene products; medications; someone to check in with; something to do; other].” Matched administrative data provided basic demographic and health information about respondents (age, gender, race, ethnicity, and presence of a mental health condition [yes; no]).

### Analysis

Descriptive statistics were used to characterize the sample, and chi-square, Fisher’s-exact, and t-tests were applied to identify significant differences by unit type (SRO vs. studio). Three separate multiple logistic regression models were used to examine predictors of perceiving risk posed by COVID-19 “very seriously” (versus less than very seriously) and engaging in handwashing and social distancing behaviors “all the time” (versus less than all the time). Covariates included age in years and bivariate indicators of increased risk due to a pre-existing condition, male gender, Black or African American race, Hispanic or Latino ethnicity, presence of a mental health condition, and living in an SRO (as opposed to a studio). Models predicting engagement in preventive behaviors additionally controlled for whether participants viewed COVID-19 health risk “very seriously.” All analyses were conducted using Stata/SE for Mac, version 16.1 (StataCorp, College Station, TX).

## Results

As noted in Table 1, 216 of 532 respondents with matched demographic data (41%) lived in SROs. Our sample was 56 years old on average (SD = 10.6), 73% male, and 61% Black or African American. Only 1% (*n* = 6) indicated that they were not aware of COVID-19 and 65% (*n* = 346) regarded it as a very serious risk to their health. Compared with those in SROs, a greater proportion of studio residents reported flu-like symptoms in the prior 30 days (6% vs. 1%, *p* = .01), increased risk due to a preexisting condition (43% vs. 32%, *p* = .01), or had a mental health diagnosis (64% vs. 53%, *p =*. *01*). About three quarters of tenants in both studios (76%) and SROs (75%) reported consistent handwashing, but significantly fewer SRO residents reported consistent social distancing (63% vs. 75%, *p* = .002). Fifty-five percent of participants said they could shelter in place for 14 days if needed. Among the 45% (*n* = 238) overall who said that they could not, significantly more studio residents reported lack of food (95% vs. 86% *p* = .013), hygiene items (81% vs. 64%, *p* = .004), and medication delivery (48% vs. 22%, *p* < .001) as factors contributing to their inability to do so.

**Table 1.** Participant Characteristics with Statistically Significant Bivariate Differences by Unit Type (Studio vs. Single-Room Occupancy)

|  | **Studio** | **SRO<sup>a</sup>** | **Total** |
| --- | --- | --- | --- |
|  | (*n* = 316) | (*n* = 216) | (*N* = 532) |
|  | *M* (*SD*)<sup>b</sup> or *n* (%) | *M* (*SD*) or *n* (%) | *M* (*SD*) or *n* (%) |
| Age (years) | 55.30 (10.53) | 57.02 (10.62) | 56.00 (10.62) |
| Gender (male) | 224 (70.9) | 163 (75.5) | 387 (72.7) |
| Race (Black or African American) | 189 (59.8) | 136 (63.0) | 325 (61.1) |
| Ethnicity (Hispanic or Latino) | 53 (16.8) | 37 (17.1) | 90 (16.9) |
| Mental health diagnosis | 202 (63.9) | 114 (52.8) | 316 (59.4)* |
| Unaware of COVID-19 outbreak<sup>c</sup> | 4 (1.3) | 2 (0.9) | 6 (1.1) |
| Views COVID-19 health risk very seriously<sup>d</sup> | 211 (66.8) | 135 (62.5) | 346 (65.0) |
| Handwashing all the time<sup>e</sup> | 239 (75.6) | 162 (75.0) | 401 (75.4) |
| Social distancing all the time<sup>f</sup> | 238 (75.3) | 136 (63.0) | 374 (70.3)** |
| Preexisting condition COVID-19 risk group<sup>g</sup> | 137 (43.4) | 70 (32.4) | 207 (38.9)* |
| Flulike symptoms in prior 30 days<sup>h</sup> | 18 (5.7) | 3 (1.4) | 21 (4.0)* |
| Can shelter in place for 14 days if needed<sup>i</sup> | 176 (55.7) | 118 (54.6) | 294 (55.3) |
| “What would you need so that you can shelter in place that you are not able to secure for yourself?”<sup>j</sup> | (*n* = 140) | (*n* = 98) | (*n* = 238) |
| Food delivery (non-perishable foods) | 133 (95.0) | 84 (85.7) | 217 (91.2)* |
| Hygiene items | 113 (80.7) | 63 (64.3) | 176 (73.9)** |
| Medication delivery | 67 (47.9) | 24 (24.5) | 91 (38.2)*** |
| Someone to check in | 46 (32.9) | 22 (22.4) | 68 (28.6) |
| Something to do | 21 (15.0) | 18 (18.4) | 39 (16.4) |
| None of the above | 3 (2.1) | 2 (2.0) | 5 (2.1) |

\*\*\* $p < .001$ , \*\* $p < .01$ , \* $p < .05$ .
<sup>a</sup> SRO = single-room occupancy unit.
<sup>b</sup> *M* = mean; *SD* = standard deviation.
<sup>c</sup> “Have you heard of the coronavirus/COVID-19 outbreak?”
<sup>d</sup> “Do you view this outbreak as a serious risk to your health?”
<sup>e</sup> “Have you been washing your hands more since the outbreak began?”
<sup>f</sup> “Have you been practicing social distancing (i.e., staying away from others and avoiding social gatherings to avoid spreading the virus)?
<sup>g</sup> “Are you in a category considered to be at serious risk for COVID-19 due to a pre-existing condition?”
<sup>h</sup> “Have you had flu-like symptoms in the past 30 days?”
<sup>i</sup> “Are you able to shelter in place (i.e., not leave your home and practice social distancing at all times) for 14 days?”
<sup>j</sup> Only includes participants who responded “no” or “maybe” to the above question. Respondents could endorse one or more options, or “none of the above.”

Regression results detailed in Table 2 indicate that older age and preexisting conditions were associated with one to two times the odds of perceiving COVID-19 to be a very serious health risk (*p* = .02 and *p* = .005, respectively). Men had significantly lower odds of perceiving risk very seriously than women (OR = 0.56, *p* = .008) Perceiving COVID-19 as a very serious health risk was associated with three to four times the odds of engaging in consistent handwashing and social distancing (*p* < .001). Those living in SROs and those with mental health conditions had about half the odds of reporting consistent social distancing (*p* = .005) and handwashing (*p* = .004) than those in studios.

**Table 2.** Results of Three Separate Multiple Logistic Regressions Examining Predictors of Viewing COVID-19 Health Risk “Very Seriously” and Consistent Engagement in Preventive Handwashing and Social Distancing Behaviors.

|  | **Very Seriously<sup>a</sup>** | **Handwashing<sup>b</sup>** | **Social Distancing<sup>b</sup>** |
| --- | --- | --- | --- |
|  | *OR (95% CI)<sup>c</sup>* | *OR (95% CI)* | *OR (95% CI)* |
| Age (years) | 1.02 (1.00, 1.04)* | 1.00 (0.98, 1.02) | 0.98 (0.96, 1.00) |
| Gender (male) | 0.56 (0.36, 0.86)** | 0.65 (0.40, 1.08) | 0.90 (0.57, 1.41) |
| Race (Black or African American) | 1.04 (0.67, 1.62) | 1.23 (0.75, 2.03) | 0.87 (0.54, 1.39) |
| Ethnicity (Hispanic or Latino) | 1.11 (0.62, 1.98) | 1.19 (0.62, 2.28) | 1.26 (0.66, 2.40) |
| Mental health diagnosis | 0.90 (0.61, 1.31) | 0.52 (0.33, 0.81)** | 0.81 (0.54, 1.21) |
| Single-room occupancy unit | 0.85 (0.59, 1.24) | 0.94 (0.62, 1.45) | 0.56 (0.38, 0.84)** |
| Preexisting condition risk group | 1.74 (1.18, 2.55)** | 0.70 (0.46, 1.08) | 0.90 (0.60, 1.36) |
| Views risk very seriously | -- | 3.32 (2.17, 5.07)*** | 2.92 (1.96, 4.36)*** |
| Constant | 0.82 (0.27, 2.51) | 4.05 (1.10, 14.87) | 6.14 (1.75, 21.56) |
| Observations | 532 | 532 | 532 |

\* $p < .05$ . \*\* $p < .01$ . \*\*\* $p < .001$ .
<sup>a</sup> 1 = “Very seriously”; 0 = “Not at all,” “not very seriously,” or “somewhat seriously.”
<sup>b</sup> 1 = “Yes, all the time”; 0 = “Not at all,” “yes, once in a while,” or “yes, sometimes.”
<sup>c</sup> *OR* = odds ratio; 95% *CI* = 95% confidence interval.

## Discussion

The results of our survey suggest PSH tenants are aware of the COVID-19 pandemic and many consider it to be a very serious health threat. Such awareness and perception were found to be strong predictors of taking protective measures (as is the case in the general population^7^). Although the majority of tenants reported taking protective measures, many reported not doing so. Our findings indicate that targeted outreach may be needed to reduce risk. For example, we found that male tenants had lower odds of perceiving COVID-19 as a serious health threat, which is consistent with prior literature.^11^ We also found that tenants with a mental health diagnosis, in particular, had lower odds of washing their hands consistently, which may indicate the need for increased mental health support and interventions that target daily functioning.

While our results are limited by reporting bias and chronological bias—attitudes and behaviors may be changing rapidly as the pandemic continues to unfold—the findings from this study also demonstrate how structural factors may influence preventive behavior. Tenants in SROs that have shared bathroom facilities had lower odds than others of social distancing. A lack of access to food, hygiene products, and medication delivery, especially among those living in studio apartments versus SROs, were common barriers to sheltering in place. While this suggests that having shared facilities may provide access to more resources, it may also reflect the higher rate of predisposing health conditions in studio residents resulting in increased need and risk of contracting the illness. This may also explain the higher reported rates of flu-like symptoms in studio residents. Regardless, PSH providers may need to plan for the delivery of supplies such as food, hygiene products, and medication in a more systematic and sustained way.

While lack of viral testing capacity may result in PSH programs only testing residents who are displaying active symptoms, a recent report from a shelter setting demonstrates that universal testing would be required to identify the high proportion of mild, pre-symptomatic, and asymptomatic cases,^12^ which are suspected to play a major role in COVID-19 transmission.^13^ The fact that PSH tenants exhibit premature aging, early onset of geriatric conditions, and require in-home supports^13^ suggests that risk of transmission within PSH may be elevated. This is particularly of concern in single-site programs, in which all tenants in the same building receive support services, as opposed to scatter-site programs, in which units are located in the community and operated by private landlords.^15^ Applying lessons learned from vulnerabilities found in nursing homes that have been described as “ground zero” for the COVID-19 pandemic,^16^ prevention and containment of outbreaks within single-site PSH will require proactive screening efforts, education of tenants and support staff (e.g., hand washing and social distancing), isolation of infected tenants, effective and nonpunitive sick leave policies for staff, and access to personal protective equipment (PPE) for in-home visits.^16^

In order to continue supportive services while maintaining social distance, PSH providers should consider options such as telehealth, which has been shown to be feasible in PSH,^17^ in addition to resources such as food delivery. Given that the duration of this pandemic is unknown, housing options including PSH will be critical to slowing the spread of the COVID-19 among those experiencing homelessness^5^ but will require ongoing disease surveillance and proactive protective measures.

In addition to possible reporting bias referenced above, limitations of this study include lack of information about how tenants first learned of COVID-19, whether they had accurate information about pre-existing conditions that puts them at risk or flu-like symptoms (as well as lack of specific mental health diagnoses), or whether they are exposed to updated information as knowledge of the pandemic increases. It is also unclear the extent to which the sample is representative of most tenants in supportive housing. While the average age and gender of our sample is similar to national estimates of the target population, the majority of our sample identified as Black or African American, which is higher than national and local estimates.^1^ Still, the fact that African Americans are disproportionately impacted by homelessness and COVID-19^18^ underlines the importance of research that can inform policies and interventions to be implemented by service providers in order to protect this vulnerable population.

## Data Availability

NA

## Acknowledgements

The authors would like to acknowledge Dr. Harmony Rhoades from USC and Dr. Randall Kuhn from UCLA for their input on early versions of the manuscript, as well as Skid Row Housing Trust for their partnership. This work was supported by the USC Center for Homelessness, Housing & Health Equity (H3E) Research.

